# The development and usability testing of two arts-based knowledge translation tools for pediatric anaphylaxis

**DOI:** 10.1101/2023.02.23.23286316

**Authors:** Shannon D. Scott, Lisa Hartling

## Abstract

Anaphylaxis, or anaphylactic reactions, are a severe allergic reaction with a rapid onset and can be fatal. Children are disproportionately at risk for hospitalization and emergency department visits due to anaphylaxis. A previously conducted mixed studies systematic review and qualitative descriptive study found that parents lacked confidence in recognizing and treating an anaphylactic reaction in their child, and were unsure of when to bring their child to the emergency department. This demonstrates that more effective knowledge translation (KT) tools are needed to satisfy parent information needs.

The purpose of this research was to work with parents to develop and test the usability of an animated video and an interactive infographic about anaphylactic reactions in children. These tools merge the best available research evidence with narratives of parent experiences to respond to their information needs. Prototypes were evaluated by parents (video n=31; infographic n=30) through usability testing in an urban emergency department waiting room in Alberta. Parents viewed the tools on an iPad and answered questions via an electronic survey. The usability survey consisted of 9, 5-point Likert items, which assessed: 1) usefulness, 2) aesthetics, 3) length, 4) relevance, and 5) future use. Parents were also asked to provide their positive and negative opinions of the tool via two free text boxes. Overall, results were positive and the tools were highly rated across most usability items. Mean scores across usability items were 4.26 to 4.71 for the video and 3.83 to 4.43 for the infographic. The scores from the usability testing suggest arts-based digital tools are useful in sharing complex health information with parents about managing an anaphylactic reaction in their child and provide meaningful guidance on how to improve KT tools to better reflect the needs of parents.

## Introduction

An anaphylactic reaction, also known as anaphylaxis, is a severe allergic reaction that happens very quickly and can be fatal (1, 2). Children are disproportionately at risk for hospitalization and emergency department visits due to anaphylaxis (3, 4). Parents have a crucial role in preventing and managing their child’s anaphylactic reactions.

Despite a rising global incidence of anaphylaxis and related hospitalization rates (5, 6), and a wide variety of written and online materials about anaphylaxis, parents experience challenges accessing this information and successfully managing the condition. Parents are not always aware of these resources, or they may feel uncertain about the credibility and reliability of the information (7, 8). They want the content of information sources to be comprehensive and evidence-based, but at the same time tailored to their reading level and information needs (8). Parental uncertainty combined with anxiety during a child’s illness are common, leading parents to seek emergency or medical care (7). The burden of anaphylaxis on families and the healthcare system highlights opportunities to reduce health system costs and improve patient and family education.

We previously conducted a mixed studies systematic review which identified that parents lacked confidence in recognizing an anaphylactic reaction in their child, felt hesitant or afraid when treating the reaction, and felt they needed more information to confidently manage their child’s anaphylaxis (9). We also conducted a qualitative descriptive study with parents who had brought their child to the emergency department during an anaphylactic reaction and found that parents reported feelings of uncertainty as to when their child needed epinephrine administered, and generally lacked confidence as to knowing if and when their child needed emergency care. The results of this study are currently being prepared for publication. Understanding these information needs is essential to creating resources that can effectively communicate health information to help parents make informed decisions when seeking health care for children experiencing an anaphylactic reaction. Furthermore, this information demonstrates that more effective knowledge translation (KT) tools are needed to satisfy parent information needs.

Research exploring the benefits of art and narrative based forms of KT tools have illustrated the power these modalities may have in communicating, engaging, and influencing individuals (10-15). Furthermore, previous research has demonstrated the positive impacts of working with end-users of health information, such as parents and other caregivers, to develop KT tools (13, 15-20). Such collaborations have resulted in tools that are relevant and meet the information needs of the appropriate stakeholders. More specifically, arts- and narrative-based KT tools have been proven to be effective sources of communication, translating complex health information into engaging and understandable content for parents (13, 15-20). Currently, there have been limited numbers of KT tools developed to provide parents education on anaphylaxis and how to manage an anaphylactic reaction in a child. The majority of the information available is in a strictly written format, or is meant to be used along with an anaphylaxis action plan that is developed in consultation with a health care provider (HCP).

Therefore, the purpose of this research was to work with parents to develop and assess the usability of an animated video and an interactive infographic about anaphylaxis and anaphylactic reactions in children. Following prototype completion and usability testing, the finalized tools were made publicly available on our website https://www.echokt.ca/tools/ and disseminated through stakeholder websites and on social media.

## Methods

A multi-method study involving patient engagement was used to develop, refine, and evaluate a whiteboard animation video and interactive infographic for a child having an anaphylactic reaction. Research ethics approval was obtained from the University of Alberta Health Research Ethics Board (Edmonton, AB) [Pro00062904]. Operational approvals were obtained from the Stollery Children’s Hospital to conduct usability testing.

### Compilation of Parents’ Narratives

We conducted a qualitative descriptive study (21-23) involving semi-structured interviews (**Appendix A**) to identify parents’ experiences and information needs and to inform parental narratives for our tools. Parents with children who presented to the Stollery Children’s Hospital Emergency Department with anaphylaxis were recruited for qualitative interviews. Parents were asked to share their experiences with having a child with the condition. The results from the qualitative descriptive study are being prepared for publication elsewhere. Concurrently, our research group conducted a mixed-studies systematic review to synthesize current evidence about experiences and information needs of parents managing anaphylaxis. Results from the systematic review (9) are published elsewhere.

### Prototype (Intervention) Development

Results from both the systematic review and qualitative interviews were used to inform the development of an infographic skeleton and video script. Clinical content from a Bottom Line Recommendation (BLR) developed by TRanslating Emergency Knowledge for Kids (TREKK) was also included in the tools (24). Following the completion of the infographic skeleton and video script, researchers worked with illustrators and graphic designers to develop the tool prototypes.

### Video

The English-language video was 5 minutes and 45 seconds long, narrated in the third person, and included closed captioning. It outlined the story of a young child named Ari who has an anaphylactic reaction. The video highlighted the process Ari took to become diagnosed with the condition and described important information about allergies, including symptoms, how anaphylaxis can be diagnosed, and how it may impact children differently. The video also outlined how allergies can be mild or severe, and what to do if a child is having a severe allergic reaction, also known as an anaphylactic reaction. The video included information about what to expect at the emergency department and what follow-up care can be expected after an anaphylactic reaction. Information is included about how to help manage a child’s anaphylaxis and what to teach a child so they can be safe. Screen captures of the video are included in **Appendix B**.

### Infographic

The interactive infographic was developed in the same format as other infographics in our suite of tools (https://www.echokt.ca/tools/). The style is unique to our research program and was developed over the course of several years (25).

The interactive infographic looks similar to a webpage and allows users to scroll through the information, exploring at their own pace. The ability for parents to control what they view based on their needs differentiates the interactive infographic from the video. The information provided in the infographic mirrors the information provided in the video and is comprised of eight major sections: (1) General Information about Anaphylactic Reactions, (2) Symptoms, (3) What To Do, (4) Treatment, (5) Follow-up, (6) Diagnosis, (7) Action Plan, (8) Preventing Anaphylactic Reactions. Within *What To Do*, the infographic provides information on what to do with epinephrine and without epinephrine, including directions on how to use three forms of autoinjectors. Within *Treatment*, the tool provides information about the three brands of epinephrine available in Canada, and describes a rebound reaction. Within *Preventing Anaphylactic Reactions*, the tool provides information about what parents can do and what they can teach their child to prevent an allergic reaction from occurring. The infographic also provides useful links to other credible websites with information about anaphylaxis in children. Screen captures of the infographic are included in **Appendix C**.

### Revisions

Iterative processes were used to develop the tools and parents, health care providers (HCPs), and researchers provided several rounds of feedback. HCPs were asked to comment on the accuracy of information and evidence. Given the evolving evidence base on treatment there was significant involvement of HCPs throughout the development of these tools. Parents from our Pediatric Parent Advisory Group (P-PAG) (25) were asked to provide feedback on the length, stylistic elements, and information not addressed in the tools. The P-PAG meets once a month and members are asked to participate in tool development several times a year. Likewise, research team meetings are held weekly to discuss the development of our tools.

### Surveys

Parents presenting with an ill child to a major pediatric emergency department (ED) in the Edmonton area were recruited to participate in an electronic, usability survey **(Appendix D)**. Members of the study team approached parents in the ED to determine interest and study eligibility. Parents who agreed to participate in the study were provided with an iPad by the researcher and asked to complete a consent form. Once the consent was completed, study participants viewed one of the digital tools (video or infographic) then were automatically directed to complete the usability survey questions. Usability testing for the video was completed prior to the infographic allowing for feedback from parents to be applied to the infographic prior to its testing. Study team members were available in the ED to provide technical assistance and answer questions as parents were completing the surveys. The usability survey included 9, 5-point Likert items that assessed: 1) usefulness, 2) aesthetics, 3) length, 4) relevance, and 5) future use. The usability survey was designed in-house based upon key elements identified by a systematic search of over 180 usability evaluations (26). Parents were also asked to provide their positive and negative opinions of the tool via two free text boxes.

### Data Analysis

Data was cleaned and analyzed using SPSS v.24. Descriptive statistics and measures of central tendency were generated for demographic questions. Likert responses were given a corresponding numerical score, with 5 being “Strongly Agree” and 1 being “Strongly Disagree” (27, 28). Means and standard deviations (SDs) were calculated for each usability item. Independent two tailed t-tests were used to determine if there was a significant difference (<0.05) in the mean usability scores of the two KT tools for each usability item. Open-ended survey data was analyzed thematically.

See **Appendix E** for an overview of the entire project timeline.

## Results

Sixty-one parents awaiting pediatric ED care completed the usability survey (30 parents participated in the infographic usability testing and 31 parents participated in the video usability testing). The demographic characteristics of the parents who participated are presented in **Table 1**.

**Table 1.** Demographic characteristics of participants who assessed the usability of the anaphylaxis digital tools (video n=30; infographic n=31; combined n=61)

| Characteristic | Infographic<br>n (%) | Video<br>(n%) | Combined<br>(n%) |
| --- | --- | --- | --- |
| Sex |  |  |  |
| Female | 26 (86.7) | 25 (80.6) | 51 (83.6) |
| Male | 4 (13.3) | 5 (16.1) | 9 (15) |
| Non-binary | 0 | 1 (3.2) | 1 (1.6) |
| Ethnicity |  |  |  |
| African American/Canadian | 2 (6.7) | 0 | 2 (3.3) |
| Asian | 2 (6.7) | 2 (6.5) | 4 (6.6) |
| Black | 1 (3.3) | 0 | 1 (1.6) |
| First Nations | 2 (6.7) | 1 (3.2) | 3 (4.9) |
| Hispanic/Latino | 0 | 1 (3.3) | 1 (3.3) |
| Metis | 0 | 2 (6.5) | 2 (6.5) |
| Middle eastern/North African | 0 | 3 (9.7) | 3 (4.9) |
| South Asian | 2 (6.7) | 2 (6.5) | 4 (6.6) |
| Southeast Asian | 2 (6.7) | 1 (3.2) | 3 (4.9) |
| White/Caucasian | 19 (63.3%) | 19 (61.3) | 38 (62.3) |
| Age |  |  |  |
| Less than 20 years old | 1 (3.3) | 1 (3.2) | 2 (3.3) |
| 20-30 years | 5 (16.7) | 4 (12.9) | 9 (14.7) |
| 31-40 years | 13 (43.3) | 19 (61.3) | 32 (52.4) |
| 41-50 years | 11 (36.7) | 6 (19.4) | 17 (27.8) |
| 51 years and older | 0 | 1 (3.2) | 1 (1.6) |
| Marital Status |  |  |  |
| Married/Partnered | 24 (80) | 22 (71) | 46 (75.4) |
| Single | 6 (20) | 9 (29) | 15 (24.6) |
| Education |  |  |  |
| Some high school | 3 (10) | 0 | 3 (4.9) |
| High school diploma | 5 (16.7) | 6 (19.4) | 11 (18) |
| Some post-secondary | 2 (6.7) | 4 (12.9) | 6 (9.8) |
| Post-secondary certificate/diploma | 2 (6.7) | 10 (32.3) | 12 (19.7) |
| Post-secondary degree | 10 (33.3) | 5 (16.1) | 15 (24.6) |
| Graduate degree | 8 (26.7) | 5 (16.1) | 13 (21.3) |
| Missing | 0 | 1 (3.2) | 1 (1.6) |
| Household Income |  |  |  |
| Less than \$25,000 | 3 (10) | 2 (6.5) | 5 (8.2) |
| \$25,000-\$49,000 | 1 (3.3) | 3 (9.7) | 4 (6.5) |
| \$50,000-\$74,000 | 5 (16.7) | 4 (12.9) | 9 (14.7) |
| \$75,000-\$99,000 | 2 (6.7) | 7 (22.6) | 9 (14.7) |
| \$100,000-\$149,000 | 4 (13.3) | 4 (12.9) | 8 (13.1) |
| \$150,000 and over | 9 (30) | 7 (22.6) | 16 (26.2) |
| Prefer not to answer | 6 (19.4) | 4 (12.9) | 10 (16.4) |
| Geographical Area |  |  |  |
| City | 22 (73.3) | 19 (61.3) | 41 (67.2) |
| Town | 1 (3.3) | 4 (12.9) | 5 (8.2) |
| Suburb | 4 (13.3) | 6 (19.4) | 10 (16.4) |
| Farm | 2 (6.7) | 1 (3.2) | 3 (4.9) |
| Other | 1 (3.3) | 1 (3.2) | 2 (3.3) |
| Relationship to the child brought to the Emergency |  |  |  |
| Parent | 27 (90) | 29 (93.5) | 56 (91.8) |
| Guardian | 1 (3.3) | 2 (6.5) | 3 (4.9) |
| Other family Member | 2 (6.7) | 0 | 2 (3.3) |
| Number of Children in the Family |  |  |  |
| 1 | 4 (13.3) | 7 (22.6) | 11 (18) |
| 2 | 12 (40) | 14 (45.2) | 26 (42.6) |
| 3 | 9 (30) | 5 (16.1) | 14 (22.9) |
| 4 | 1 (3.3) | 4 (12.9) | 5 (8.2) |
| 5+ | 2 (6.7) | 2 (3.2) | 4 (6.5) |
| Missing | 2 (6.7) | 0 | 1 (1.7) |
| Times Visited ED |  |  |  |
| 1-5 Times | 19 (63.3) | 20 (64.5) | 39 (63.9) |
| 6+ Times | 10 (33.3) | 11 (35.5) | 21 (34.4) |
| Missing | 1 (3.3) | 0 | 1 (1.7) |

Overall, the tools were rated highly by parents with most selecting strongly agree or agree for the usability items **(Table 2)**. The mean scores across usability items for the video ranged from 4.26 to 4.71 (on a 5-point scale with 4 indicating agree and 5 indicating strongly agree). Parents felt the video was useful and relevant to them. They found the video easy to use and felt it could be used without written instruction. Parents also felt the length of the video was appropriate and thought it was aesthetically pleasing. Parents agreed and strongly agreed that they would use the video in the future and that it would help them make decisions about their child’s health. Finally, when asked if they would recommend the video to a friend, parents agreed or strongly agreed.

**Table 2.** Means (SD) of participant responses to the usability survey.

| Usability Measures | Video | Infographic | P-value |
| --- | --- | --- | --- |
| It is useful. | 4.55 (0.50) | 4.43 (0.62) | 0.499 |
| It provides information that is relevant to me as a parent. | 4.39 (0.91) | 4.30 (0.65) | 0.713 |
| It is simple to use. | 4.71 (0.46) | 4.27 (0.64) | 0.002* |
| I can use it without written instructions or additional help. | 4.58 (0.62) | 3.83 (0.87) | <0.001* |
| Its length is appropriate. | 4.35 (0.71) | 4.03 (0.73) | 0.090 |
| It is aesthetically pleasing (i.e. images, colours, etc.). | 4.35 (0.60) | 4.14 (0.83) | 0.340 |
| It helps me make decisions about my child's health. | 4.32 (0.70) | 4.07 (0.75) | 0.182 |
| I would use it in the future. | 4.26 (0.73) | 4.07 (0.75) | 0.184 |
| I would recommend it to a friend. | 4.45 (0.67) | 4.10 (0.77) | 0.030* |

\*p value of <0.05 is considered significant.

For the infographic, mean scores across usability items ranged from 3.83 to 4.43 (on a 5-point scale with 3 indicating neither agree nor disagree and 4 indicating agree). Parents felt that the infographic was useful and relevant to them as parents. When parents were asked whether the infographic was simple to use, parents agreed and strongly agreed, however, parents were either neutral or agreed when asked if they felt that the tool could be used without written instructions or additional help. Parents felt that the length of the infographic was appropriate and that it was aesthetically pleasing. Parents agreed that they would use the infographic in the future and that it would help them make decisions about their child’s health. Finally, when asked if they would recommend the infographic to a friend, parents agreed.

There was a statistically significant difference between mean ratings for the video versus the infographic on three of the usability items. Differences for two items had to due with the simplicity of the tool to use and whether parents felt they could use it without written instructions or additional assistance. Specifically, parents felt the video was simpler to use than the infographic and that they could use it without written instructions or additional help. Parents were also less likely to recommend the infographic to their friends when compared to the video. No significant differences were found between the two tools in terms of usefulness, relevance of information to them as a parent, length of the tool, aesthetics of the tool, the ability of the tool to help parents in decision making about their child’s health, and the likeliness the parent would use it in the future.

There were a few negative comments in the open text responses about the video, two were about the video being “a bit too long” and a couple of others provided suggestions on additional content such as “explain what epinephrine is” and how it works by saying “blue to the sky, orange to the thigh”. There were many positive comments about the video: parents described it as “simple and easy to understand”, “easy to follow” and “engaging”. There were a few negative comments about the infographic, one being that a user-centered interaction “wasn’t obvious”, and one being that there was “too much medical jargon”. There were many positive comments: parents described the infographic as “clear”, “very easy to use”, “easy to read”, and “visually pleasing”.

As a result of the suggestions from usability testing, additional information was added to the infographic about epinephrine and how it works. Because the video was already considered long, this information was not added to the video. As usability testing for the video was completed prior to usability testing of the infographic, this information was contained in the version of the infographic that was tested. As a result of the suggestions for the infographic about the user-centered interaction not being obvious, the final version of the tool was modified to make the functions more noticeable. Further, the language in some areas was changed to be easier to understand (i.e., plain language).

## Conclusions

The purpose of this project was to develop two arts-based digital tools (an animated video and interactive infographic) for parents of children who experience anaphylactic reactions. Using a multi-method approach that encompassed stakeholder engagement, we created tools that were highly rated amongst parents seeking care for their children in an urban emergency care centre. Parents found the tools to be useful, relevant, easy to use, and aesthetically pleasing. Most importantly, parents felt that the tools could facilitate decision-making in the future, with parents mainly agreeing and strongly agreeing that they would use the tool and recommend the tool to their friends. Of interest is that parents assessed the video as more usable than the infographic for three items. Important differences between usability items related to the simplicity of the tool and whether it could be used without written instructions or additional help. These results are insightful in terms of how parents perceive the usability of different modalities. Overall, the results from this project indicate that end-user engagement plays a positive role in developing knowledge tools that address the needs of parents.

### The tools can be found here: <u>echokt.ca/anaphylaxis</u>

Note: Our KT tools are assessed for alignment with current, best-available evidence every two years. If recommendations have changed, appropriate modifications are made to our tools to ensure that they are up-to-date (29).

## Other Outputs from this Project

### Research Papers

Rahman S, Elliott SA, Scott SD, Hartling L. Children at risk of anaphylaxis: A mixed-studies systematic review of parents’ experiences and information needs. PEC Innovation. 2022; 1:100018. doi: 10.1016/j.pecinn.2022.100018

### Presentations & Research Conferences

Sun, C., Ashry, M.E., Lubimiv, S., Knisley, L., Scott, S.D. Evaluating Social Media Metric Changes on Instagram to Disseminate an Anaphylaxis Tool to the General Public: A Repeated Measures Feasibility Study. Poster presentation. Women and Children’s Health Research Institute (WCHRI) Research Day. Edmonton, AB. November 2, 2022.

## Supporting information

Supplemental Table 1: Adapted Reporting Checklist for Multi-Method research

## Data Availability

All data produced in the present study are available upon reasonable request to the authors.

https://www.echokt.ca/anaphylaxis

## Author Contributions

This study was conducted under the supervision of Dr. Shannon D. Scott (SDS) and Dr. Lisa Hartling (LH), principal investigators (PIs) for **translating Evidence in Child Health to enhance Outcomes** (ECHO) Research and the **Alberta Research Centre for Health Evidence** (ARCHE), respectively. Both PIs designed the research study and obtained research funding through the Canadian Institutes of Health Research (CIHR).

SDS designed and supervised all aspects of tool development and evaluation.

LH designed and provided input on all aspects of tool development and evaluation.

All authors contributed to the writing of this technical report and provided substantial feedback.

## Acknowledgments

Given COVID-19 pandemic restrictions which prevented ECHO research staff from collecting the usability data, University of Alberta Pediatric Emergency Medicine research staff were contracted to collect usability data.

Anne Le (AL) conducted and analyzed qualitative interviews with parents.

Kathy Reid (KR) analyzed the usability data.

## This work was funded by

### Canadian Institutes of Health Research

- Scott, S.D. (co-PI) & Hartling, L. (co-PI), Ali, S., Currie, G., Dyson, M., Fernandes, R., Fleck, B., Freedman, S., Jabbour, M., Johnson, D., Junker, A., Klassen, T., Maynard, D., Newton, A., Plint, A., Richer, L., Robinson, J., Robson, K., Vandall-Walker, V. [all collaborators listed in alphabetical order]. (2016) Integrating evidence and parent engagement to optimize children’s healthcare. CIHR Foundation Scheme ($2,500,000). July 2016-June 2023.

## Appendices

## Appendix A Qualitative Interview Guide

Parents will be interviewed to understand their experience having a child with an anaphylactic reaction. Semi-structured interviews will be conducted with parents in order to get their “narrative” or experiences. The following questions will be used to guide these interviews. Being true to semi-structured interview techniques, interview questions will start broad and then move to the more specific.

1. Tell me about your experience with your child having an anaphylactic reaction
  a. Tell me about the events <u>leading up</u> to your child having an anaphylactic reaction.
    i. What was your child doing? (eating? Did they take a medication? Were they bit/stung by an insect? exposed to something?)
    ii. Signs/symptoms your child was experiencing (Child’s health – skin rash, flushing, trouble breathing, vomiting, etc.)
    iii. Were there other factors happening at the <u>same time</u> as exposure to the allergen that could have influenced the reaction? (for instance, was the child exercising, were they ill? Were they taking any medications? Was your child under an increased emotional stress? Or recently traveled outside the country?).
    iv. Things you tried to decrease the symptoms (remove the cause of the reaction, EpiPen, administer Benadryl)
    v. How did <u>your child feel</u> during this time? (emotions, mental state)
    vi. How did <u>you feel</u> during this time? (emotions, mental state)
    vii. When and how did you decide to go to the ED?
2. Was this the first time your kid has had an anaphylactic reaction?
  a. If not, tell me about when you child had their first anaphylactic reaction (or how you determined they could have an anaphylactic reaction).
  b. Confirm child’s age when diagnosed with allergies and their first anaphylactic reaction. How did you find out your child had allergies? Does your child have any other illnesses (e.g. Asthma, cystic fibrosis, etc.)?
  c. What happened <u>after</u> your child had their first anaphylactic reaction?
    i. Medications ordered? (tell me about the medications they take and response to medications)
    ii. Education given for day-to-day management
    iii. Follow up care provided?
    iv. Were there changes that the family needed to make for the child with the allergy/ies? (Probe – removing allergens from the home, etc.)
3. Tell me about the <u>day-to-day management of your child’s allergies (prevention of anaphylaxis)</u>
  a. Probe for a sense of are the child’s allergies (anaphylaxis prevention) well managed or are their frequent reactions?
  b. How did you feel about managing your child’s allergies (day to-day) when they were first diagnosed– Probe for parent’s level of comfort/knowledge/skill being able to manage their child’s allergies.
    i. Are there particular challenges? For instance, sending your kid to school/daycare, stigma associated with always having an EpiPen, sleepovers/playdates, playing sports, etc. (attempting to “control” that environment)
    ii. How do you feel now about managing your child’s allergies (probing if there has been an increasing of knowledge over time)?
  c. Did you feel that you had adequate knowledge, know-how to manage your child’s allergies when they were first diagnosed? How about now?
    i. Probe where they referred to an Allergy Specialist?
    ii. Did they get the Canadian Anaphylaxis Action Plan for Kids guide?
    iii. Was their child prescribed an EpiPen or other medications?
  d. If you went to the ED with your child’s anaphylactic reaction, tell me about that experience (if there are many – suggest the last experience, or the most memorable experience).
    i. What assessments were done
    ii. Medications
    iii. How was your child during the experience (emotions)?
    iv. How were you during the experience
  e. Has having allergies affected other aspects of your child’s life?
    i. School attendance
    ii. Friends/Social Support
4. Are there “things” that make it challenging to care for your child with allergies?
  a. Probe for – knowledge needs? (If yes, what would they like to know more about); is it parental knowledge needs or the need for others in their child’s live to know more about allergies (e.g., teachers, coaches, grandparents).
  b. Environmental challenges (where they live, where the child goes to school, weather/seasonal challenges)
  c. Personal challenges (the costs associated with having a child with allergies – e.g. cost of EpiPen, etc.)

## Appendix B Images from video about anaphylaxis in children (video available at https://www.echokt.ca/anaphylaxis/)

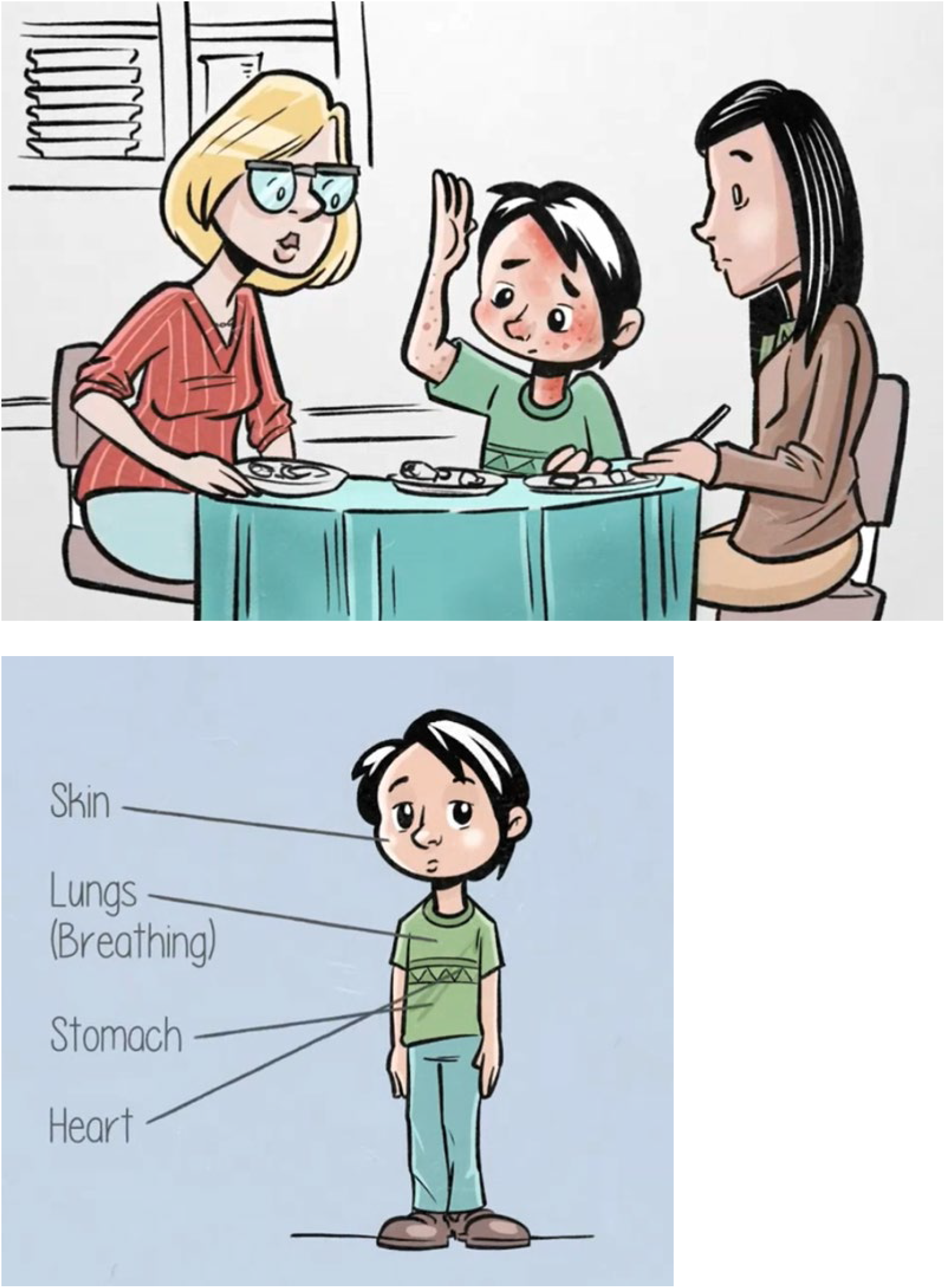

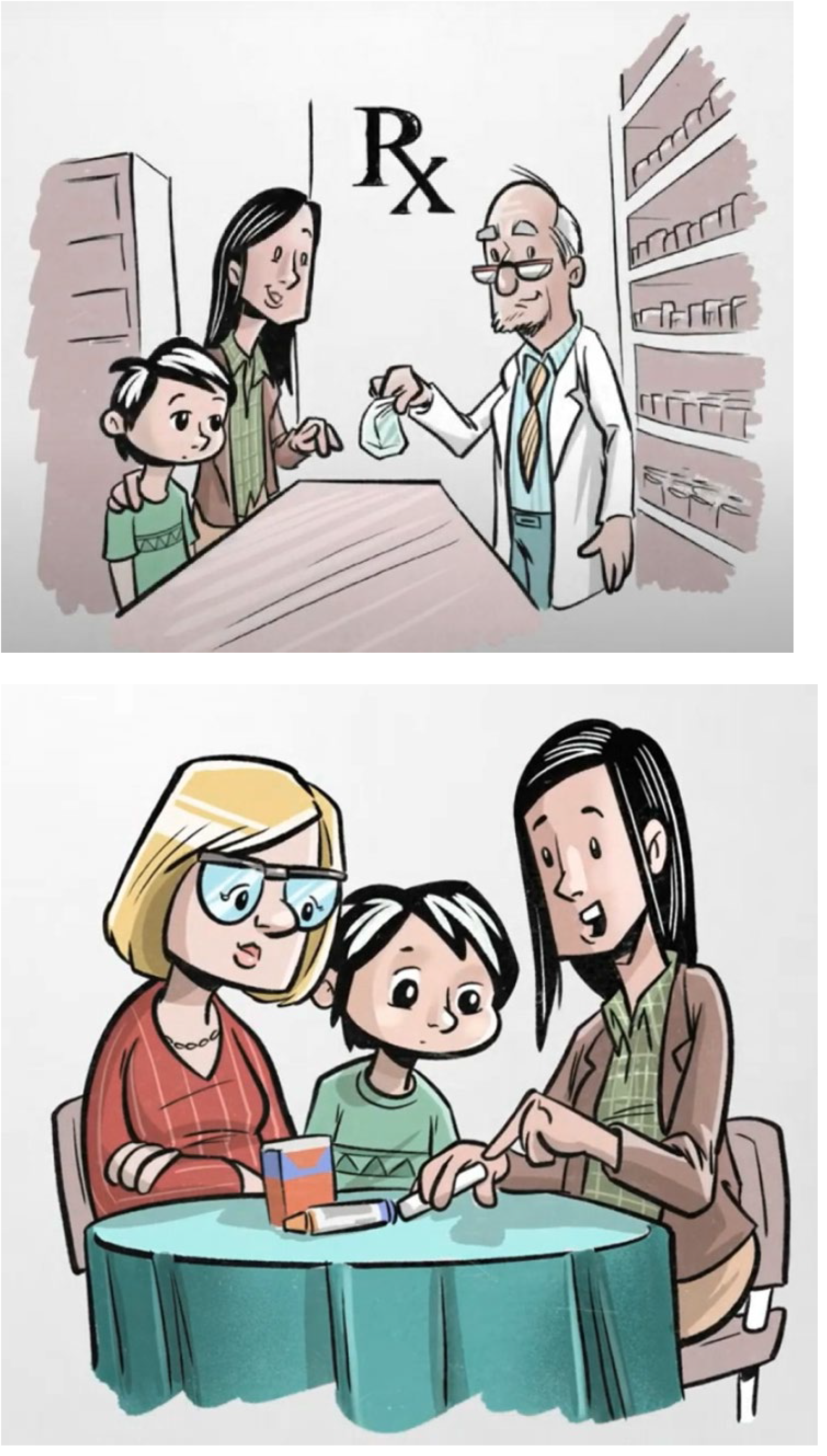

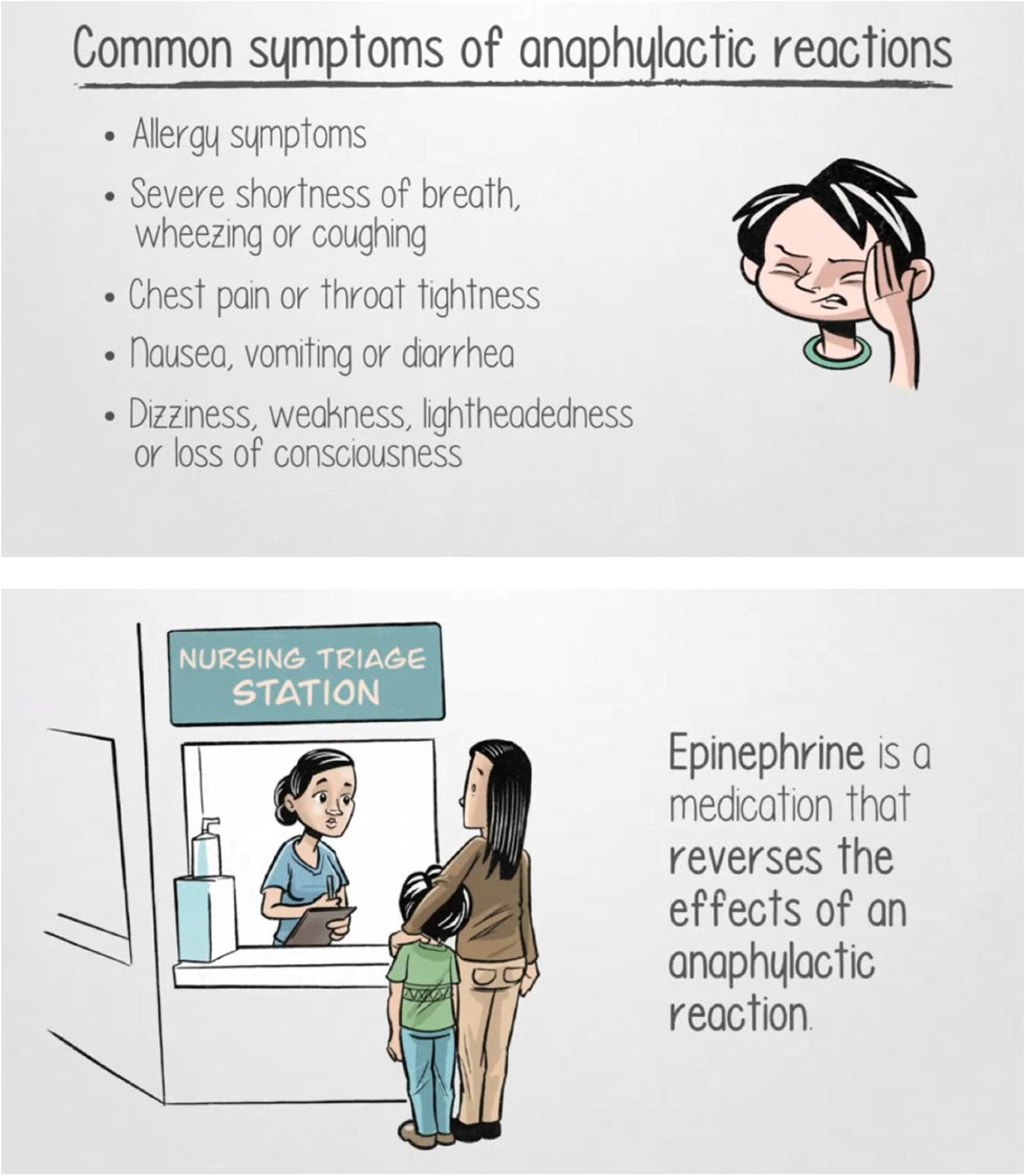

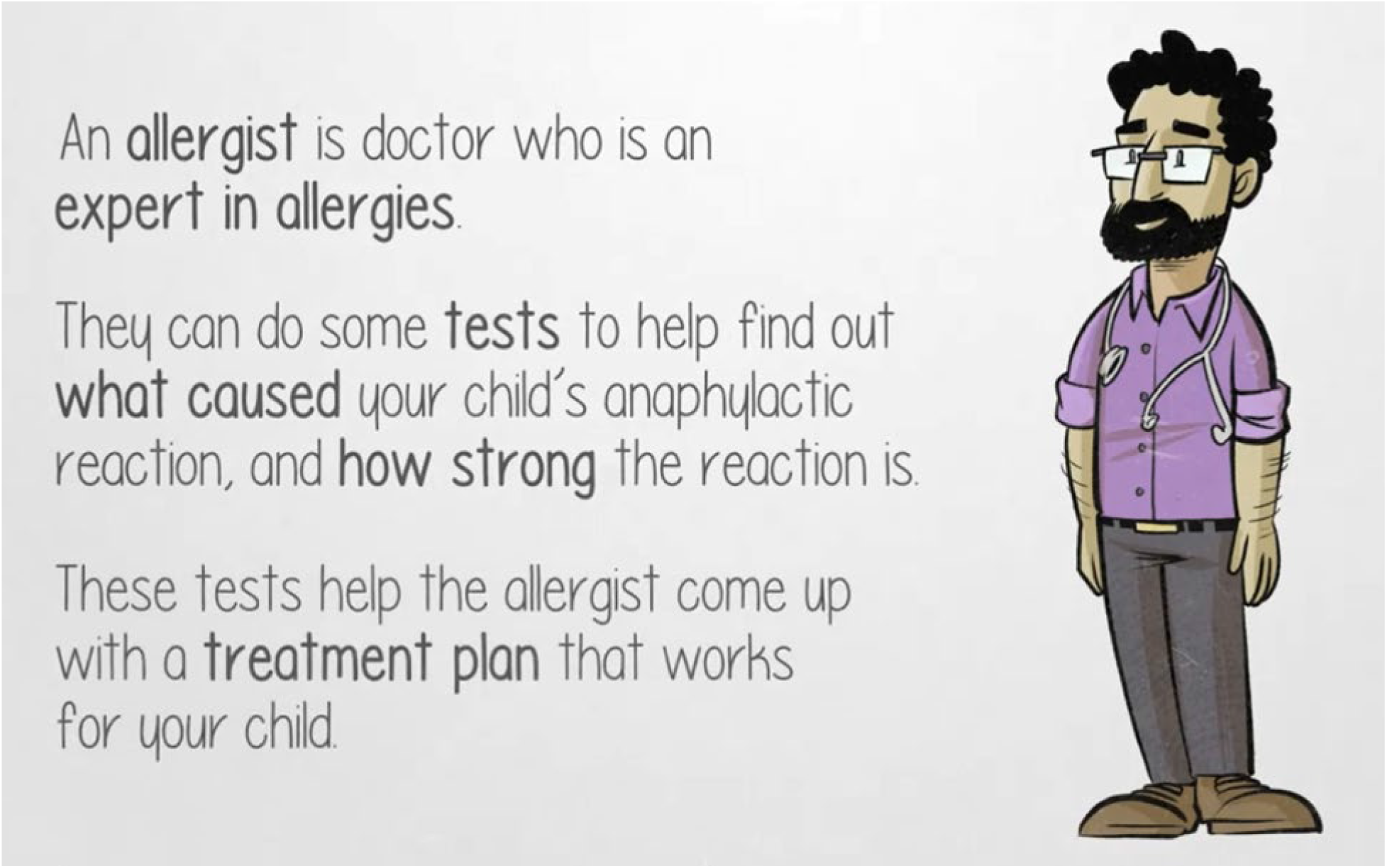

## Appendix C Images from infographic about childhood asthma (infographic available at https://www.echokt.ca/anaphylaxis-infographic/)

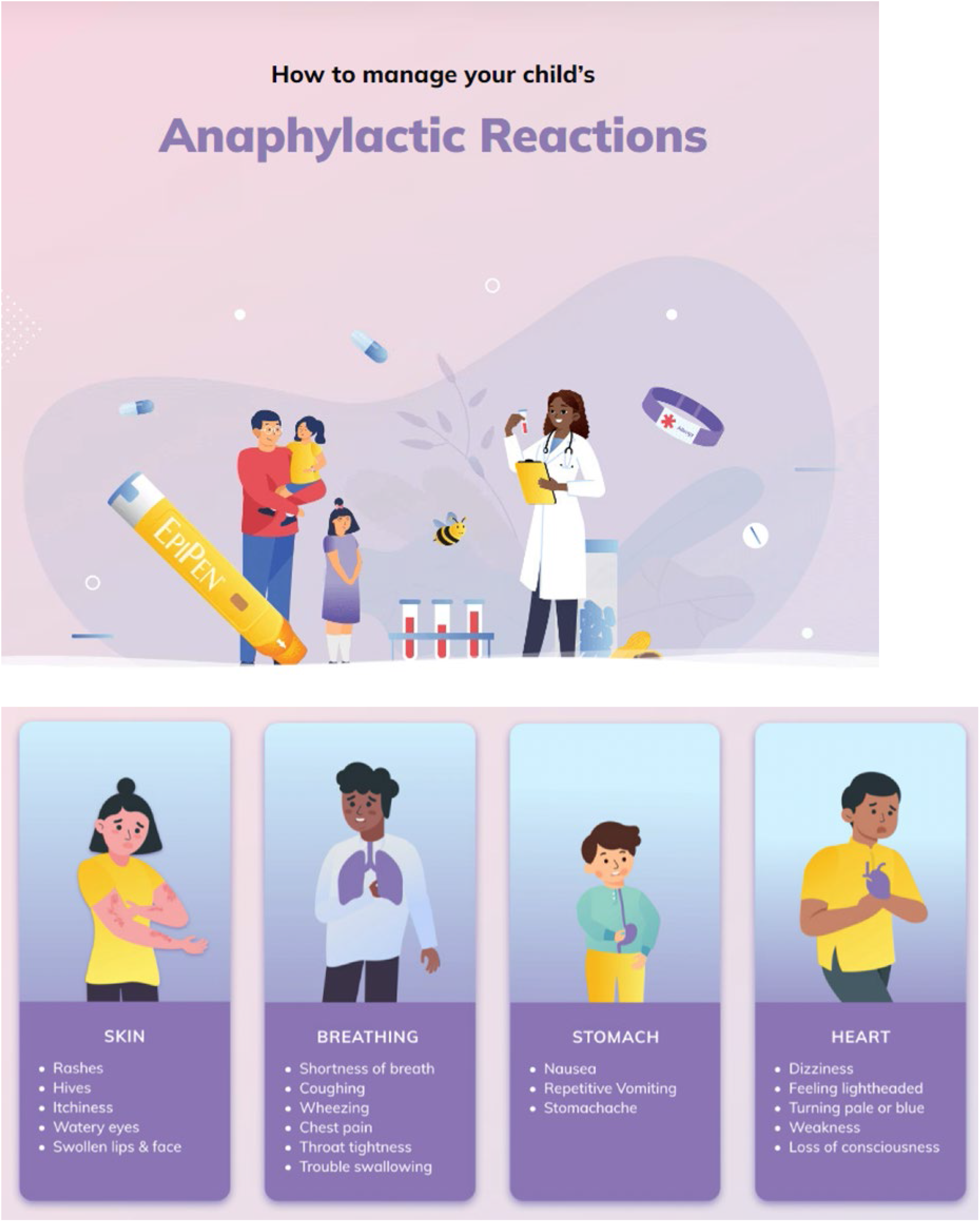

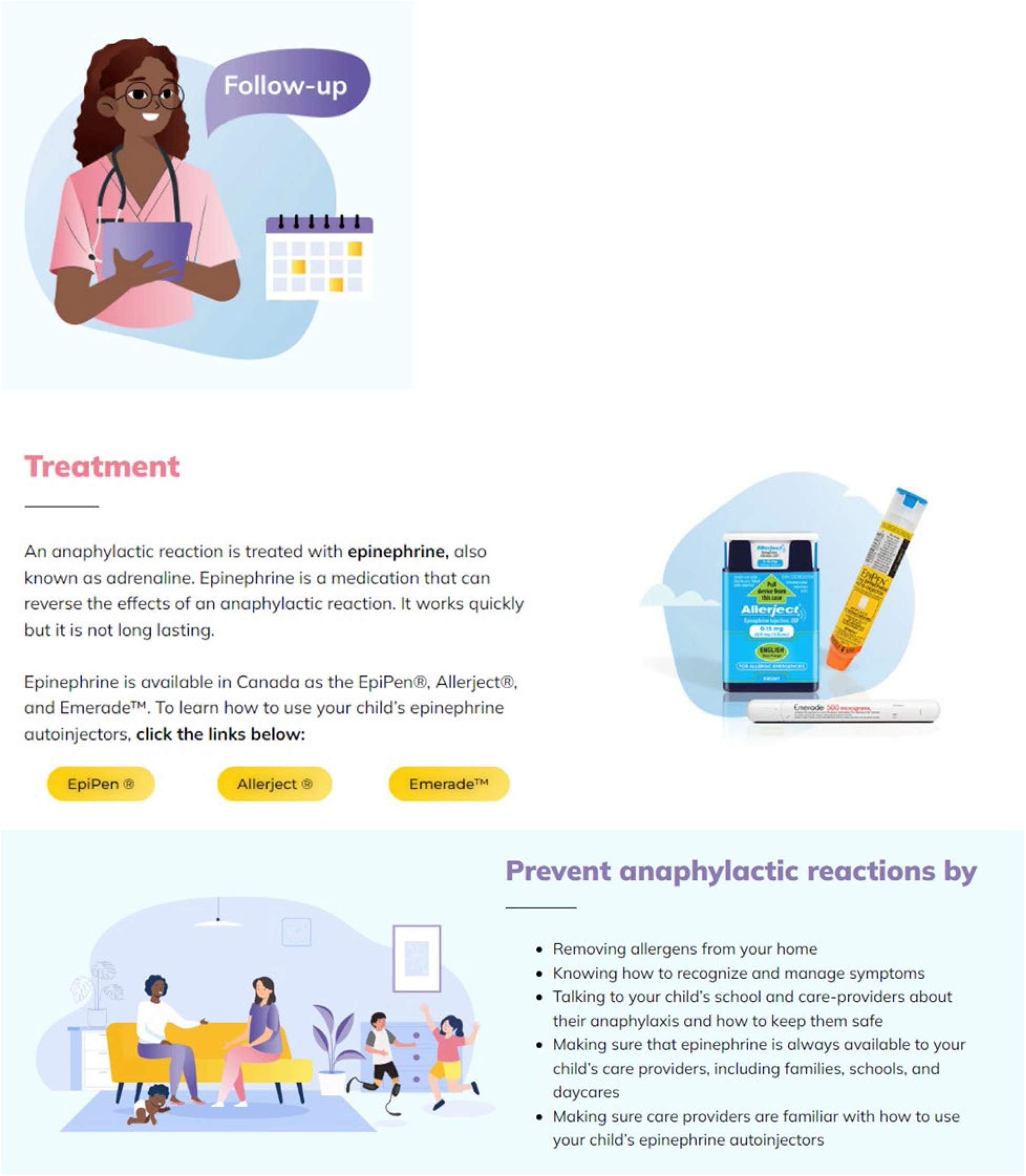

## Appendix D Usability Survey

SECTION 1: Demographics

1. a. What is your gender?
  □ Male
  □ Female
  □ Non-binary
  □ Two-spirit
  □ Other: _____________
  □ Prefer not to answer

1. b. Would you describe yourself as transgender?
  □ Yes
  □ No
  □ Prefer not to answer
2. Which ethnicities best describes you? *Please select all that apply*.
  □ Asian
  □ African American or African Canadian
  □ Black
  □ First Nations
  □ Hispanic or Latino
  □ Métis
  □ Middle Eastern or North African
  □ South Asian
  □ Southeast Asian
  □ White or Caucasian
  □ Not listed: _____________
  □ Prefer not to answer
3. What is your Age?
  □ Less than 20 years old
  □ 20-30 years
  □ 31-40 years
  □ 41-50 years
  □ 51 years and older
4. What is your Marital Status?
  □ Married/Partnered
  □ Single
5. What is your gross annual household income?
  □ Less than $25,000
  □ $25,000-$49,999
  □ $50,000-$74,999
  □ $75,000-$99,999
  □ $100,000-$149,999
  □ $150,000 and over
  □ Prefer not to answer
6. What is your highest level of education?
  □ Some high school
  □ High school diploma
  □ Some post-secondary
  □ Post-secondary certificate/diploma
  □ Post-secondary degree
  □ Graduate degree
  □ Other: _____________
7. Where does your household live
  □ City
  □ Suburb
  □ Town
  □ Farm
  □ Other: _____________
8. What is your relationship to the child that you have brought to the emergency department?
  □ Parent
  □ Grandparent
  □ Other family member
  □ Guardian
9. How many children do you have?
10. How old are your children?
11. How many times have you visited the emergency department with your children?
  □ 1-5 times
  □ 6+times
12. Have any of your children ever been admitted to the hospital?
  □ Yes
  □ No

SECTION 2: Assessment of attributes of the arts-based, digital tools

Note: items 1-9 are rated on a 5-point Likert scale from 1=strongly disagree to 5=strongly agree

1. It is useful. [5-point Likert Scale]
2. It provides information that is relevant to me as a parent. [5-point Likert Scale]
3. It is simple to use. [5-point Likert Scale]
4. I can use it without written instructions or additional help. [5-point Likert Scale]
5. Its length is appropriate. [5-point Likert Scale]
6. It is aesthetically pleasing (i.e., images, colours, etc.). [5-point Likert Scale]
7. It helps me to make decisions about my child’s health. [5-point Likert Scale]
8. I would use it in the future. [5-point Likert Scale]
9. I would recommend it to a friend. [5-point Likert Scale]
10. List the most positive aspects: [open text]
11. List the most negative aspects: [open text]

## Appendix E Project Timeline

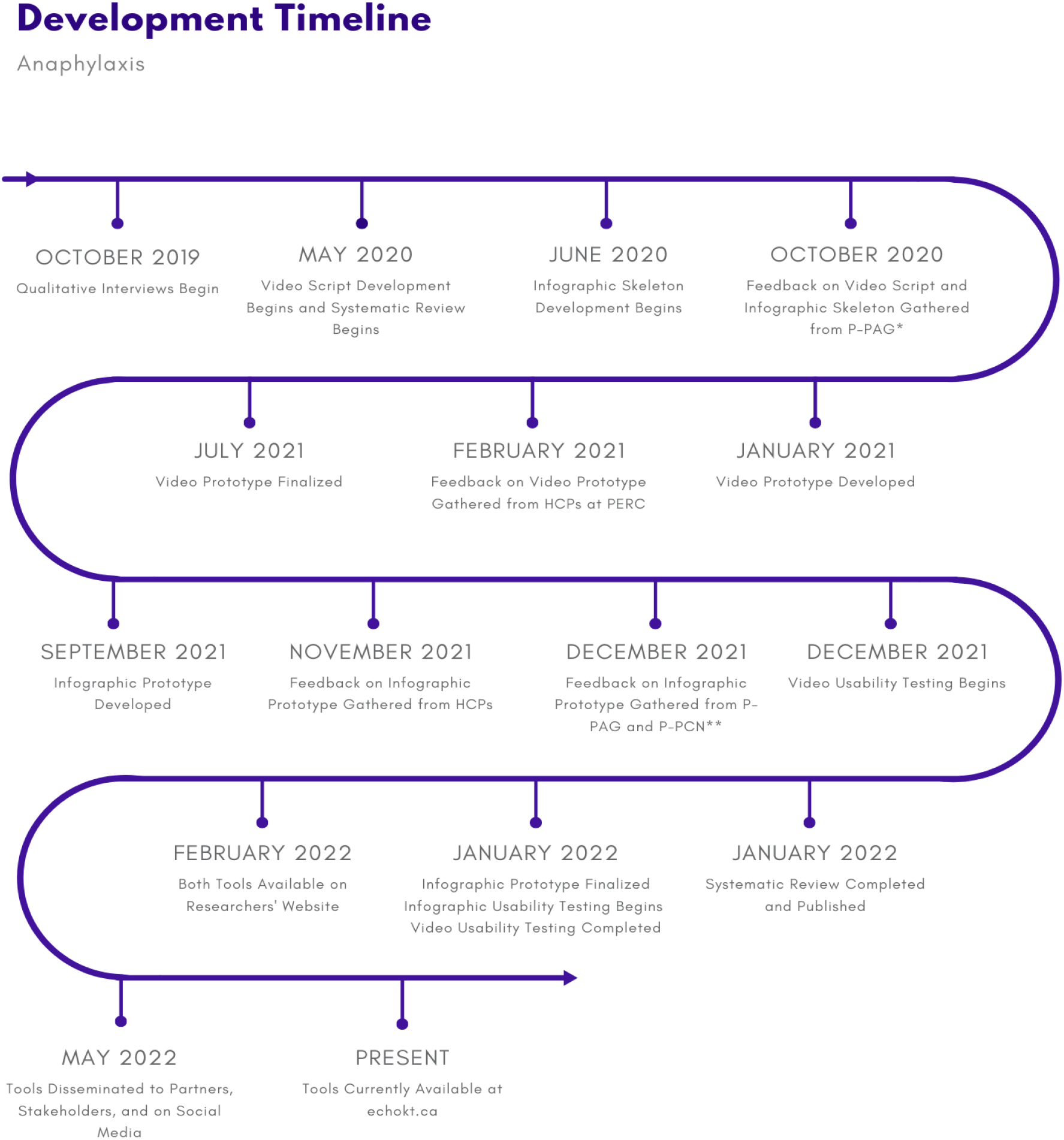

* P-PAG = Pediatric Parents’ Advisory Group (P-PAG)

** P-PCN = Pediatric Parent Consultation Network

HCPs = Health Care Providers

PERC = Pediatric Emergency Research Canada

